# Computational characterization of lymphocyte topology on whole slide images of glomerular diseases

**DOI:** 10.1101/2025.04.12.25325548

**Authors:** Xiang Li, Manav Shah, Qian Liu, Jin Zhou, Gina Sotolongo, Jeffrey B. Hodgin, Laura Mariani, Lawrence Holzman, Andrew R. Janowczyk, Jarcy Zee, Kyle J. Lafata, Laura Barisoni

## Abstract

The complexity of distribution of inflammatory cells in the kidney is not well captured by conventional semiquantitative visual assessment. This study aims to computationally quantify the topology of lymphocytic inflammation and tested its clinical relevance.

N=333 NEPTUNE/CureGN participants (N=155 focal segmental glomerulosclerosis (FSGS) and N=178 Minimal Change Disease (MCD) with available clinical/demographic data and 1 Hematoxylin & Eosin-stained whole slide image (WSI), were included. Deep learning models were applied to segment cortex and lymphocytes. Graph modeling, where nodes were defined as lymphocytes and edges as the spatial connections between cortical lymphocytes, were applied to all WSIs. We then developed a novel graph-based habitat clustering algorithm to identify dense vs. sparse lymphocytic habitats. From each habitat, 26 high-throughput quantitative pathomic features were extracted to capture cell density, connectivity, clustering, and centrality.

The association of these pathomic features with disease progression (40% eGFR decline or kidney replacement therapy) was assessed using LASSO-regularized Cox proportional hazards models. Clinical and demographic characteristics were added as potential confounders. Kaplan-Meier survival analysis with log-rank test was used to evaluate risk stratification. Two validation strategies were applied: (i) training on NEPTUNE with external validation on CureGN data, and (ii) using an 80/20 data partition of the combined datasets for training and validation, respectively.

Multivariable Cox models integrating clinical/demographic variables with graph features achieved validation concordance index of 0.736±0.072 in the CureGN external validation and 0.757±0.071 in the combined validation dataset. The average degree feature (overall connectivity) in dense habitat and k-core feature (clustering pattern strength) in sparse habitat revealed consistent association with clinical outcome.

The topological characterization of lymphocytic inflammation identifies immune habits, capturing the complexity of pattern of inflammation beyond human vision. These pathomic/topology signatures represent potential digital biomarkers that can enhance our ability to prognosticate/predict clinical outcome in MCD/FSGS.

## Introduction

The complex interplay between immune system dysregulation and kidney disease progression has emerged as a critical area of research in nephrology^1–3^. Investigations into biomarkers such as the neutrophil-to-lymphocyte ratio^4^ and relative lymphocyte count^5^ have shown promise in predicting chronic kidney disease (CKD) progression and kidney failure. Similarly, the severity of interstitial inflammation has also been proven to be associated with disease progression in glomerular diseases^6^, although its role in nephrotic syndrome is understudied. Pathologists characterize interstitial inflammation based on its (i) composition (lymphocytes, plasma cells, neutrophils, eosinophils, etc), (ii) whether it is limited to the interstitium or is spatially associated with kidney structures (glomerulitis, peritubular capillaritis, tubulitis, arteritis), and (iii) its severity using a semiquantitative scoring approach (none, mild, moderate, and severe). However, interstitial inflammation can be sparse, dense, focal, diffuse, or a combination of all the above. Such complexity in the distribution of inflammatory cells cannot be well captured by the human eye nor represented by a semiquantitative/ gestalt approach.

The advent of digital pathology has revolutionized the field of pathology, allowing for the digitization of entire tissue sections at high resolution and the development of computational methods to accurately quantify tissue characteristics and discover new digital biomarkers for kidney diseases^7^. Deep learning (DL) approaches have shown promise in segmentation and efficient analysis of histologic structures in the kidney^8–18^. However, the spatial distribution of inflammatory cells in the kidney and its clinical relevance remain under-explored.

In this study, we leveraged computational pathology (cell graph and cell theory) and the infrastructure and datasets of large longitudinal studies such as the NEPhrotic syndrome sTUdy NEtwork (NEPTUNE) and Cure Glomerulonephropathy (CureGN) to study the clinical relevance of topological patterns of inflammation in proteinuric diseases such as focal segmental glomerulosclerosis (FSGS) and minimal change disease (MCD).

## Material and methods

### Disease cohorts

The study utilized data from the NEPTUNE^19^ and Cure CureGN^20^ consortia: multi-site observational cohort studies of children and adults with glomerular diseases, enrolled at the time of or within 5 years after their first clinically indicated kidney biopsy, respectively. Inclusion criteria used in this study are the following: (1) a digital renal biopsy with at least 1 Hematoxylin & Eosin (H&E) whole slide image (WSI) in the NEPTUNE/CureGN digital pathology repository; (2) a diagnosis of MCD or FSGS; (3) available clinical and demographic information; and (4) enrollment time from biopsy less than 6 months for CureGN participants.

### Demographic and Clinical data

Demographic data included age, sex, and race. Clinical data included estimated glomerular filtration rate (eGFR) and urine protein creatinine ratio (UPCR) at the time of the biopsy and during follow-up, kidney replacement therapy (chronic dialysis or kidney transplant) during follow-up, and immunosuppression treatment records at the time of the biopsy. The clinical outcome of interest was disease progression defined by the time from biopsy to a 40% eGFR decline or kidney failure (eGFR <15 mL/min/1.73m^2^ or kidney replacement therapy).

### WSI dataset and visual assessment

H&E WSIs scanned at 40x magnification went through quality control using HistoQC^21^ to exclude poor quality images. For NEPTUNE participants, the percent of interstitial fibrosis and inflammation was prevously assessed by NEPTUNE pathologists and retrieved from the NEPTUNE study dataset, while for CureGN, digital biopsies were assessed by study pathologists for percent of cortex involved by interstitial fibrosis and inflammation.

### Cortex segmentation

First, we developed a Dense-Net model^22^ to automatically segment cortex. N = 10 manually annotated H&E WSIs were used as initial ground truth label for cortical regions. Images were processed at a 256x256 patch size under 5x magnification, maintaining an 80/20 training and testing case split at WSI level. Preliminary cortex segmentation produced by the model were iteratively refined through a feedback loop with pathologists, using a total of 50 WSIs and QuPath^23^. The model was then applied to all WSIs and results were reviewed for accuracy.

### Lymphocyte segmentation

A previously developed and published DL model for the automatic segmentation of lymphocytes was applied to all NEPTUNE/CureGN H&E WSIs^24^. (Figure 1.1) Cells within the segmented cortical areas were retained and used to construct cell graphs.

**Figure 1.**
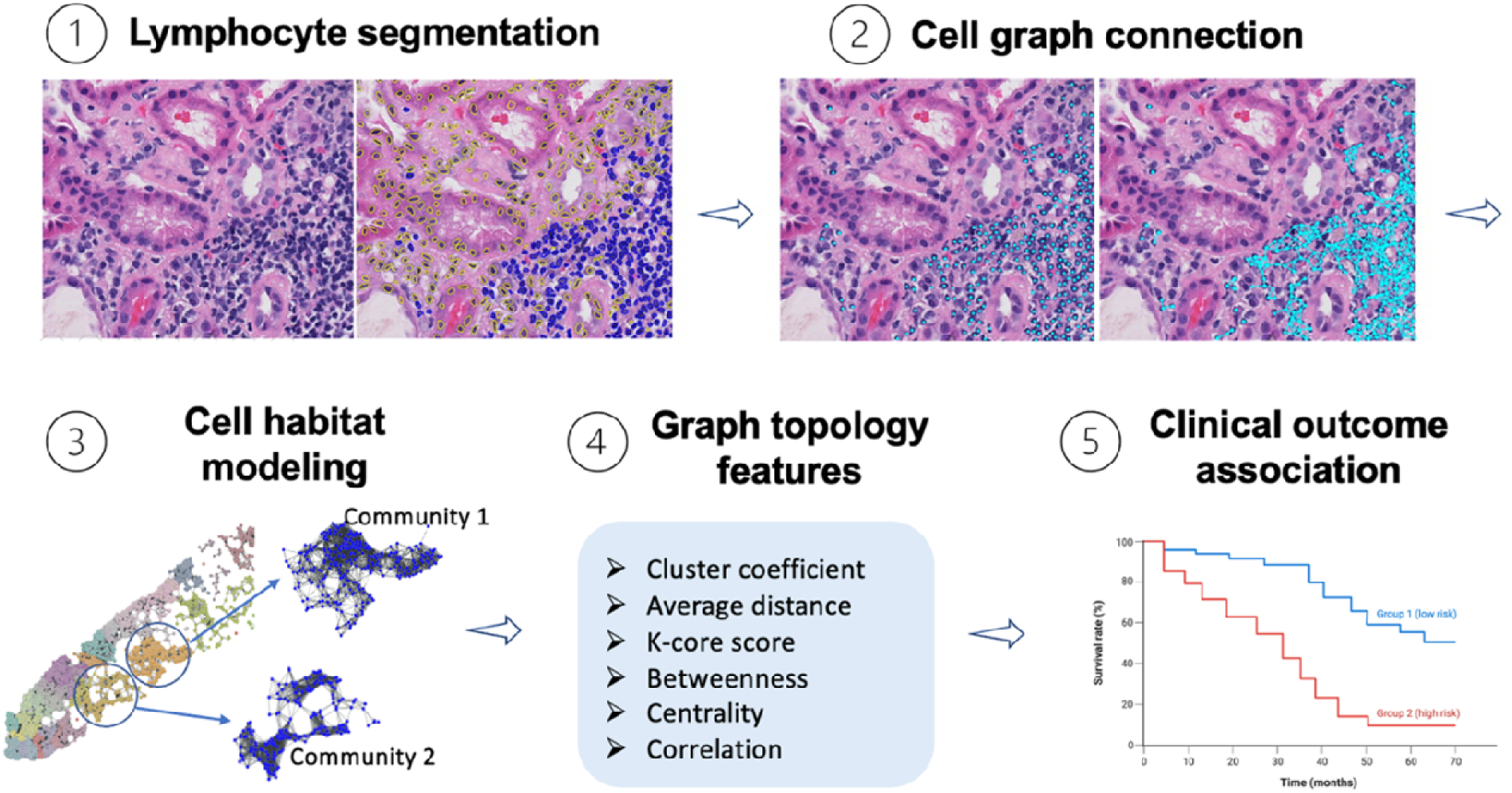
Overview of the computational framework for the characterization of lymphocyte topology on whole slide images. (1) The first step of this process involved the automatic segmentation of lymphocytes. Segmented lymphocytes were used to construct cell graphs (2) and communities (3), from which graph topology-based pathomic features were extracted (4) to describe lymphocyte spatial distribution. (5) Graph topology features were then used to test their association with clinical outcome.

### Lymphocyte graph topology

Cell graph theory was used to study the spatial relationships between lymphocytes in the tissue. Each lymphocyte was viewed as a graph node and the spatial connections between the lymphocytes was defined as graph edge. Using the cell graphs and nodes and cell graph modeling, we designed a hierarchical framework to identify cell habitats that represent the pattern of spatial distribution of lymphocytes within the cortex. Each cell habitat was defined as a group of cells that share common spatial patterns of clustering (dense habitat) or diffusing (sparse habitat). This was accomplished through the following steps: (1) lymphocytes which were within 25 µm from each other were connected with graph edges to define the local cellular neighborhood^25^ (Figure 1.2); (2) local graph habitats were identified by decomposing the whole slide graph into small groups of connected cell nodes that had similar connectivity patterns^26^. Similar connectivity patterns occurs when the number of connections within 25 µm between the cells/nodes is the same across multiple cells/nodes; (3) a total of 13 quantitative graph features were derived within each local lymphocyte group to describe graph patterns (i.e. first order, topology, correlation, clustering, and centrality) (Supplemental Table 1); (4) these local graph habitats were clustered into 2 global graph habitats using a K-means algorithm based on their graph features, which we defined as dense vs. sparse inflammation. Local graph habitats belonging to dense habitats had higher connectivity (High number of cells are within a 25 µm distance from each other) than those belonging to sparse habitats (Low number of cells are within a 25 µm distance from each other). (Figure 1.3); (5) from each of these global habitats (e.g., dense vs. sparse), we aggregated the 13 quantitative graph features by computing the mean values across all constituent local habitats, yielding a comprehensive set of 26 quantitative graph (pathomic) features that characterized the two global habitats (Figure 1.4). A complete list of graph features used in the analysis is provided in the Supplemental Table 1 as defined by graph-tool library^27^.

### Association between graph topology features and clinical outcome

(Figure 1.5) To assess the association between graph topology features and clinical outcome, we conducted domain shift (i.e. covariate shift) characterization and univariate and multivariate survival analysis. All graph features and clinico-demographic covariates were pre-processed with z-score normalization.

First, we characterized the effect of potential domain shift between NEPTUNE and CureGN cohorts through a progressive domain mixture experiment. A fraction of CureGN data (10 cases) was progressively added into NEPTUNE data to generate training datasets with increasing domain heterogeneity. A Cox proportional hazards regression model used the full feature set (including clinico-demographic information, visual scores and graph topology features) to predict clinical outcome. Model performance was evaluated using concordance index (C-index) as a function of amount of CureGN data integration, with testing performed on the remaining CureGN cohort.

Second, we tested the association between individual graph features and outcome using the NEPTUNE dataset and feature-specific univariate Cox regression models. To access whether graph features add value to the clinical management of glomerular diseases, we adjusted each univariate model by incorporating clinico-demographic information as potential confounders. Performance was compared between the feature-specific univariate model (i.e., *unadjusted*) and the feature-specific multivariate model (i.e., *adjusted*).

Third, we developed multivariable survival models consisting of *multiple* graph features as well as clinic-demographic information and visual scores. A LASSO-regularized Cox proportional hazards regression was performed to identify the most discriminative predictors. Model performance was based on the C-index. Risk stratification of high-risk vs. low-risk cohorts was defined based on median risk score calculated from the model and characterized via Kaplan-Meier analysis and log-rank test. Model robustness was assessed via bootstrap resampling (100 permutations) and validated through two experimental designs: (1) NEPTUNE for training with CureGN for external validation, and (2) combined cohorts with random 80:20 partitioning for training and validation, respectively.

## Results

### Patient Characteristics

A total of N=218 NEPTUNE (119 FSGS and 99 MCD) and N=115 CureGN (61 FSGS and 54 MCD) cases with at least 1 H&E WSI that passed quality control and with available demographic, clinical outcome, and visual assessment for interstitial fibrosis and inflammation were included (Table 1). Cases with less than 5 lymphocyte count were excluded from building graphs. Cell graphs were available in 196/218 NEPTUNE H&E WSIs (107 FSGS and 89 MCD) and 90/115 CureGN H&E WSIs (48 FSGS and 42 MCD). N =39 and N=17 disease progression events were observed from NEPTUNE and CureGN cases respectively.

**Table 1.** Patient characteristics from NEPTUNE and CureGN datasets, including clinical variables, demographic variables, and visual scoring variables.

|  |  | NEPTUNE | CureGN |
| --- | --- | --- | --- |
| Sex (%) | Male | 58% | 60% |
|  | Female | 42% | 40% |
| Hispanic (%) | No | 74% | 84% |
|  | Yes | 26% | 16% |
| Race (%) | Others | 72% | 74% |
|  | Black | 28% | 26% |
| Cohort (%) | FSGS | 55% | 53% |
|  | MCD | 45% | 47% |
| Immunosuppression (%) | No | 72% | 61% |
|  | Yes | 28% | 39% |
| Age (%) | Pediatric | 47% | 34% |
|  | Adult | 53% | 66% |
| Age (IQR) |  | 29 (11-46) | 35 (2-76) |
| UPCR at Biopsy (IQR) |  | 6.3 (1.0-7.8) | 5.8 (1.3-8.2) |
| eGFR at Biopsy (IQR) |  | 83.4 (58.7-105.3) | 81.0 (59.1-105.4) |
| Interstitial Fibrosis (IQR) |  | 12.5 (0.0-14.8) | 16.0 (0.0-25.0) |
| % Inflammation score (IQR) |  | 9.6 (0.0-14.8) | 12.3 (0.0-18.8) |
| Progression outcome event |  | 39 | 17 |
| * FSGS = focal segmental glomerulosclerosis |  |  |  |
| * UPCR = urine protein-to-creatinine ratio |  |  |  |
| * MCD = minimal change disease |  |  |  |
| * eGFR = estimated glomerular filtration rate |  |  |  |
| *Immunosuppression is at biopsy or within 30 days before biopsy |  |  |  |

### Characterization of domain shift between datasets

The domain shift (i.e. covariate shift) investigation showed a major domain gap between datasets (Figure 2). When trained exclusively on NEPTUNE data (0% CureGN inclusion), the model showed a performance degradation of 0.076 units in the C-index when applied to the CureGN validation data. As CureGN representation in the training set increased incrementally (11%, 22%, 33%, 44%, 56%, and 67%), we observed three distinct trends: (1) a slightly decreased performance from 0.826 to 0.813 for the mixed training set as domain heterogeneity increased, (2) a consistent improvement in CureGN validation performance from 0.750 to 0.835 in the C-index, and (3) an increased validation performance variability (as measured by standard deviation), attributable to the diminishing sample size in the validation set. Notably, at approximately 33% CureGN inclusion in the training set, model training and validation both reached a balanced performance of approximately 0.82 in the C-index.

**Figure 2.**
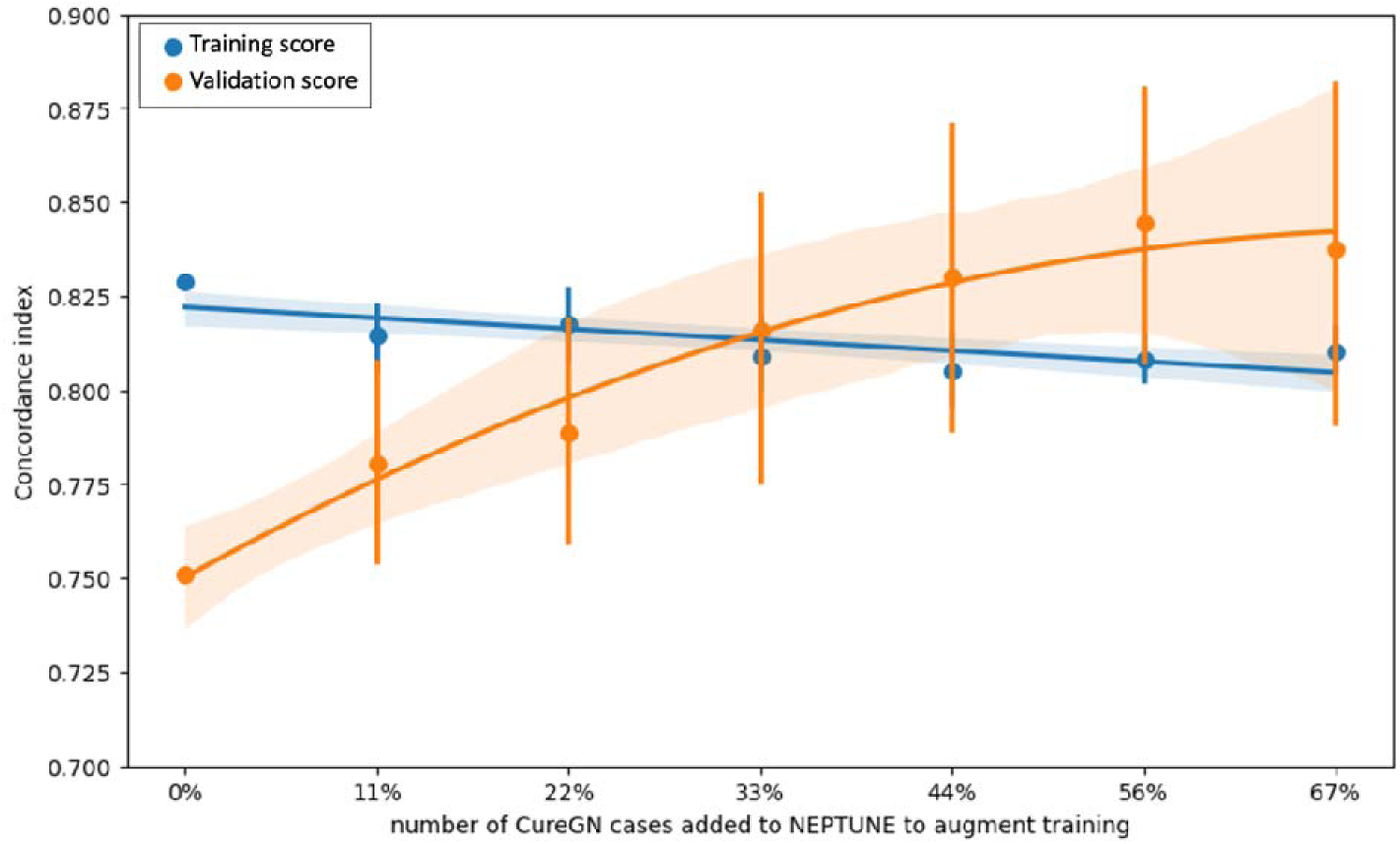
Illustration of the impact of domain shift on model performance. The x-axis represents the percentage of CureGN cases added (0-67%), while the y-axis shows the concordance index. Blue and orange lines indicate fitted training and validation performance trend with a standard deviation from 100 permutation tests, respectively. As CureGN data is incorporated into NEPTUNE, the model validation performance on the remaining CureGN data improves, with a balanced training and validation performance at 33% CureGN inclusion.

### Feature-specific modeling of disease progression

Feature-specific Cox regression models assessed the independent prognostic value across lymphocyte graph topology features from both sparse and dense habitats (Table 2). In the unadjusted univariate analyses, multiple graph features demonstrated significant associations with disease progression (p<0.05). The strongest associations based on magnitudes of hazard ratios from sparse and dense habitats were observed for *Diameter* from sparse habitat (HR = 1.39; 95% CI: 1.14-1.70; p = 0.001) and *Mean K-core* from dense habitat (HR = 1.40; 95% CI: 1.16-1.69; p < 0.001), with each HR representing a 1-SD increase in the respective feature. After adjustment for clinico-demographic characteristics, most associations were attenuated. The feature, *Average Degree* extracted from dense lymphocyte habitats, maintained independent prognostic value (HR = 1.22; 95% CI: 1.01-1.47; p = 0.043).

**Table 2.**
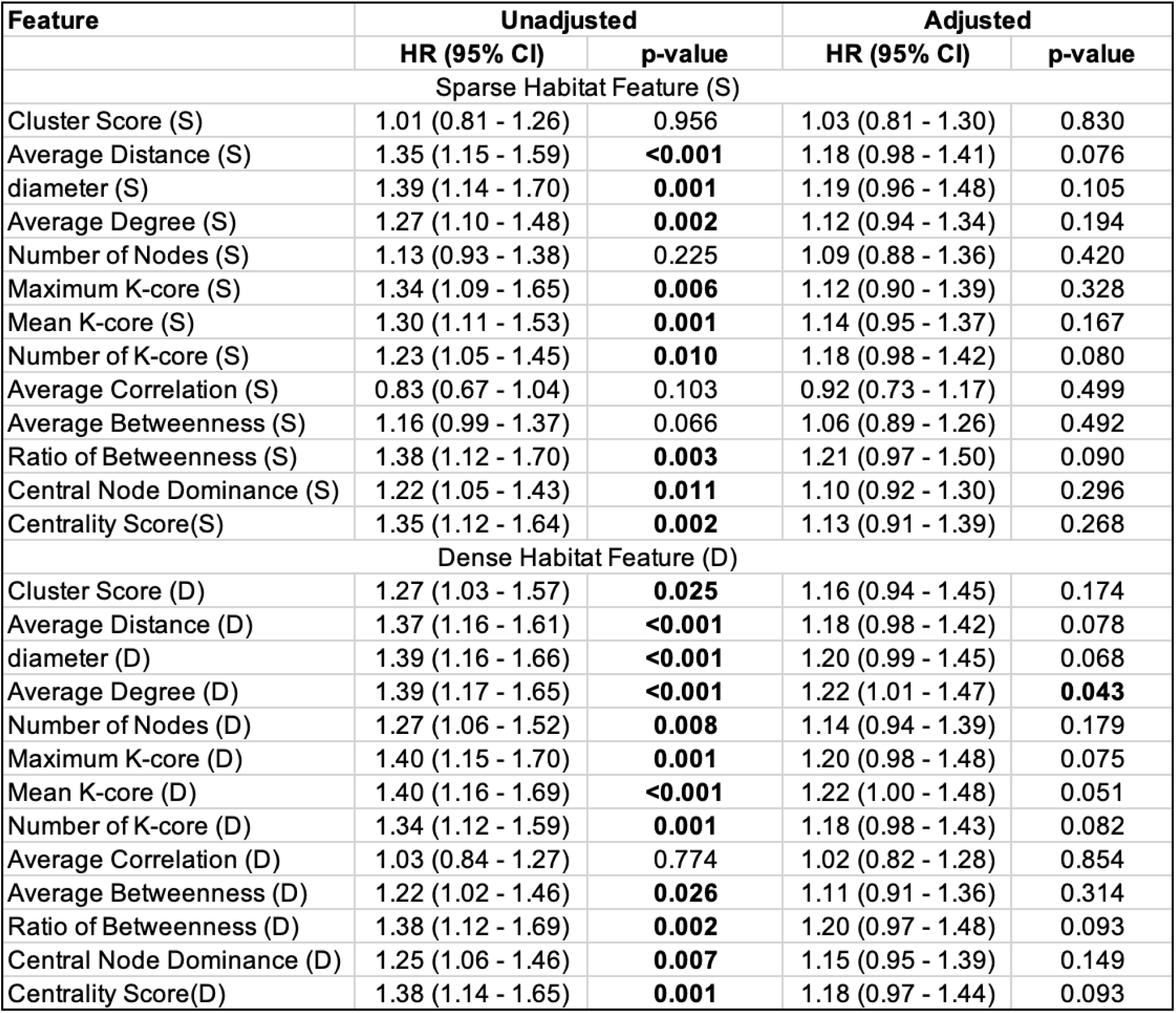
Feature-specific Cox regression model analysis for graph features. Feature *Average Degree* in dense habitat remained significant with HR = 1.22 (95% CI: 1.01-1.47), p = 0.043 after clinical indicator adjustment.

| Feature | Unadjusted |  | Adjusted |  |
| --- | --- | --- | --- | --- |
|  | HR (95% CI) | p-value | HR (95% CI) | p-value |
| Sparse Habitat Feature (S) |  |  |  |  |
| Cluster Score (S) | 1.01 (0.81 - 1.26) | 0.956 | 1.03 (0.81 - 1.30) | 0.830 |
| Average Distance (S) | 1.35 (1.15 - 1.59) | **<0.001** | 1.18 (0.98 - 1.41) | 0.076 |
| diameter (S) | 1.39 (1.14 - 1.70) | **0.001** | 1.19 (0.96 - 1.48) | 0.105 |
| Average Degree (S) | 1.27 (1.10 - 1.48) | **0.002** | 1.12 (0.94 - 1.34) | 0.194 |
| Number of Nodes (S) | 1.13 (0.93 - 1.38) | 0.225 | 1.09 (0.88 - 1.36) | 0.420 |
| Maximum K-core (S) | 1.34 (1.09 - 1.65) | **0.006** | 1.12 (0.90 - 1.39) | 0.328 |
| Mean K-core (S) | 1.30 (1.11 - 1.53) | **0.001** | 1.14 (0.95 - 1.37) | 0.167 |
| Number of K-core (S) | 1.23 (1.05 - 1.45) | **0.010** | 1.18 (0.98 - 1.42) | 0.080 |
| Average Correlation (S) | 0.83 (0.67 - 1.04) | 0.103 | 0.92 (0.73 - 1.17) | 0.499 |
| Average Betweenness (S) | 1.16 (0.99 - 1.37) | 0.066 | 1.06 (0.89 - 1.26) | 0.492 |
| Ratio of Betweenness (S) | 1.38 (1.12 - 1.70) | **0.003** | 1.21 (0.97 - 1.50) | 0.090 |
| Central Node Dominance (S) | 1.22 (1.05 - 1.43) | **0.011** | 1.10 (0.92 - 1.30) | 0.296 |
| Centrality Score(S) | 1.35 (1.12 - 1.64) | **0.002** | 1.13 (0.91 - 1.39) | 0.268 |
| Dense Habitat Feature (D) |  |  |  |  |
| Cluster Score (D) | 1.27 (1.03 - 1.57) | **0.025** | 1.16 (0.94 - 1.45) | 0.174 |
| Average Distance (D) | 1.37 (1.16 - 1.61) | **<0.001** | 1.18 (0.98 - 1.42) | 0.078 |
| diameter (D) | 1.39 (1.16 - 1.66) | **<0.001** | 1.20 (0.99 - 1.45) | 0.068 |
| Average Degree (D) | 1.39 (1.17 - 1.65) | **<0.001** | 1.22 (1.01 - 1.47) | **0.043** |
| Number of Nodes (D) | 1.27 (1.06 - 1.52) | **0.008** | 1.14 (0.94 - 1.39) | 0.179 |
| Maximum K-core (D) | 1.40 (1.15 - 1.70) | **0.001** | 1.20 (0.98 - 1.48) | 0.075 |
| Mean K-core (D) | 1.40 (1.16 - 1.69) | **<0.001** | 1.22 (1.00 - 1.48) | 0.051 |
| Number of K-core (D) | 1.34 (1.12 - 1.59) | **0.001** | 1.18 (0.98 - 1.43) | 0.082 |
| Average Correlation (D) | 1.03 (0.84 - 1.27) | 0.774 | 1.02 (0.82 - 1.28) | 0.854 |
| Average Betweenness (D) | 1.22 (1.02 - 1.46) | **0.026** | 1.11 (0.91 - 1.36) | 0.314 |
| Ratio of Betweenness (D) | 1.38 (1.12 - 1.69) | **0.002** | 1.20 (0.97 - 1.48) | 0.093 |
| Central Node Dominance (D) | 1.25 (1.06 - 1.46) | **0.007** | 1.15 (0.95 - 1.39) | 0.149 |
| Centrality Score(D) | 1.38 (1.14 - 1.65) | **0.001** | 1.18 (0.97 - 1.44) | 0.093 |

### Multivariable modeling of disease progression

The multivariable Cox survival model performance for different data partitioning strategies is shown in Table 3. When trained exclusively on NEPTUNE data and validated on CureGN data, the model exhibited high training performance (C-index = 0.864±0.030) but reduced validation performance (C-index = 0.736±0.072). In contrast, when using a mixed cohort approach (80% of combined NEPTUNE and CureGN data for training and 20% for validation), we observed more balanced performance metrics with training C-index of 0.822±0.015 and validation C-index of 0.757±0.071. Kaplan-Meier curves were used to visualize the stratified high-risk and low-risk patient groups in Figure 3. In both data partition settings, the models achieved substantial separation between patient groups for fitting and validation datasets.

**Figure 3.**
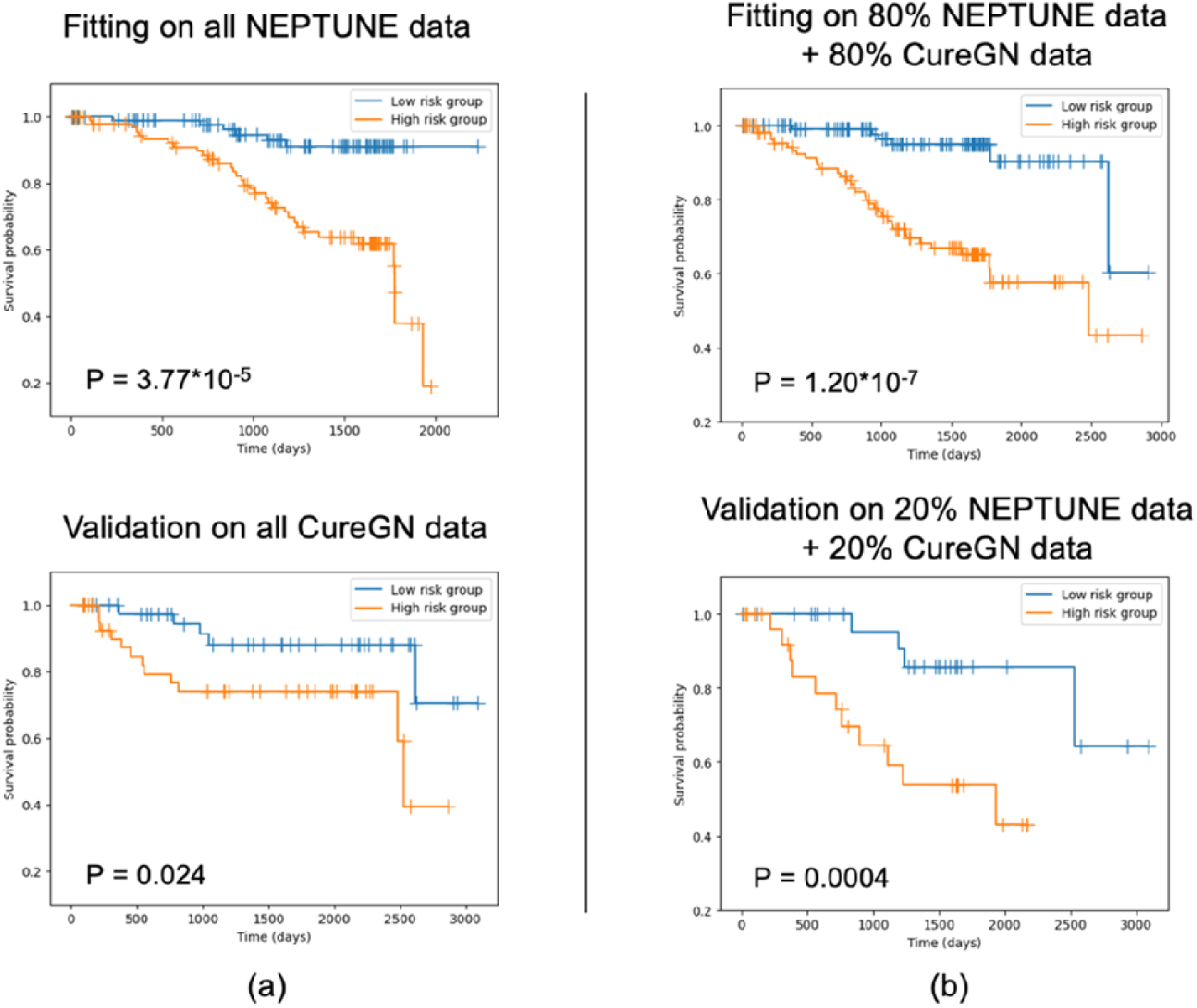
Kaplan-Meier curves of progression outcome under different data partition settings. High-risk vs. low-risk patient groups were defined based on median risk score for each situation.

**Table 3.** Multivariate Cox model performance for the progression outcome under different data partition settings. The two settings include (a) Using NEPTUNE as training dataset, and CureGN as validation dataset; (b) Using 80% NEPTUNE+CureGN as training dataset and remaining as validation dataset.

| Data split settings<br>(training : validation) | Training performance<br>(C-index) | Validation performance<br>(C-index) |
| --- | --- | --- |
| 100% NEPTUNE:<br>100% CureGN | 0.864±0.030 | 0.736±0.072 |
| 80% (NEPTUNE+CureGN) :<br>20% (NEPTUNE+CureGN) | 0.822±0.015 | 0.757±0.071 |

The LASSO-regularized feature selection process quantified feature stability through selection frequency distribution across 100 bootstrap permutations (Figure 4a). The frequency distribution revealed that clinical and pathology features - *Diagnosis* (FSGS vs. MCD) and *Interstitial Fibrosis* (visual assessment) – were consistently identified as robust predictors, being selected in >90% of permutations. Among graph topology features, several demonstrated notable selection frequency. When we studied the distribution of the feature coefficient for each selected feature across the same permutations (Figure 4b), we observed that the *Average Degree* feature in dense habitat increased risk, and *max k-core*num k-core* feature in sparse habitat decreased risk. An example of visualization of global dense and sparse habitat formulation process, and of graph feature values mapped with specific graph topology patterns is shown in Figure 5. Notably, cases with similar semiquantitative visual score for interstitial inflammation have different feature values for dense and sparse cell habitat (Figure 6).

**Figure 4.**
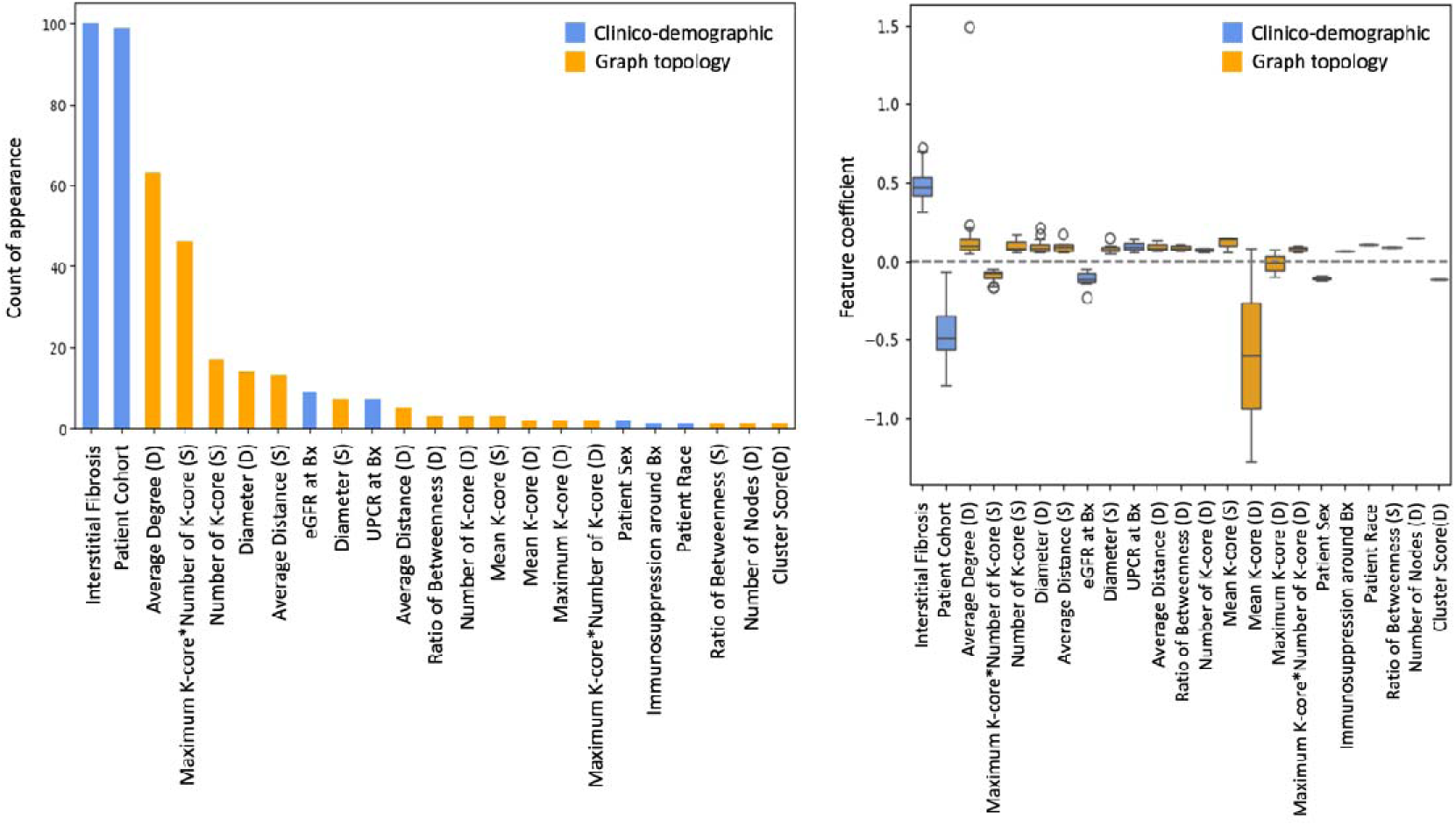
Feature selection from LASSO-regularized Cox models. (a) Frequency plot of each feature being selected in 100 permutations of Cox models with LASSO regularization. (b) Boxplot of feature coefficients for each feature being selected from the same permutation process. For both figures, blue color indicates clinico-demographic features, and orange color indicates graph features.

**Figure 5.**
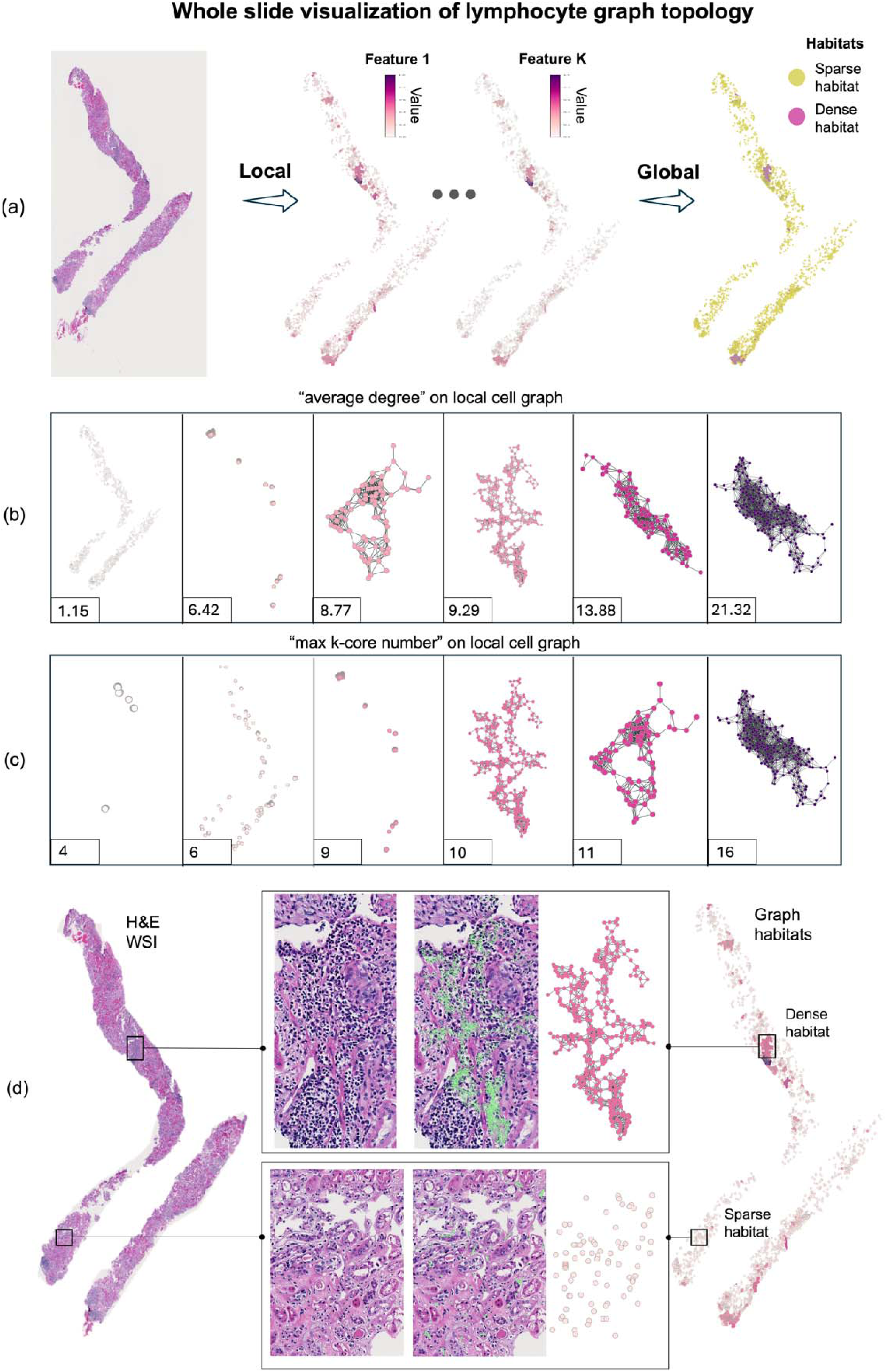
Graph feature visualization on an example WSI case. (a) Global habitats derived from all local habitats. There are two dense habitat regions in this case, while the remaining portions of cortex are occupied by sparse habitats. (b)-(c) Examples of graph habitat patterns. For the selected graph features – *average degree* (measure of how many connections nodes typically have) and *max kcore* (measure of the most densely clustered graph patterns), different feature values (i.e. the numbers shown on bottom left corner) are related to different graph topology patterns. The numbers at the bottom indicate increasing average degree / max k-core number in the graphs. (d) H&E WSI showing dense inflammation region (upper box) and sparse inflammation region (lower box) with the same region overlay by local cell graph habitat.

**Figure 6.**
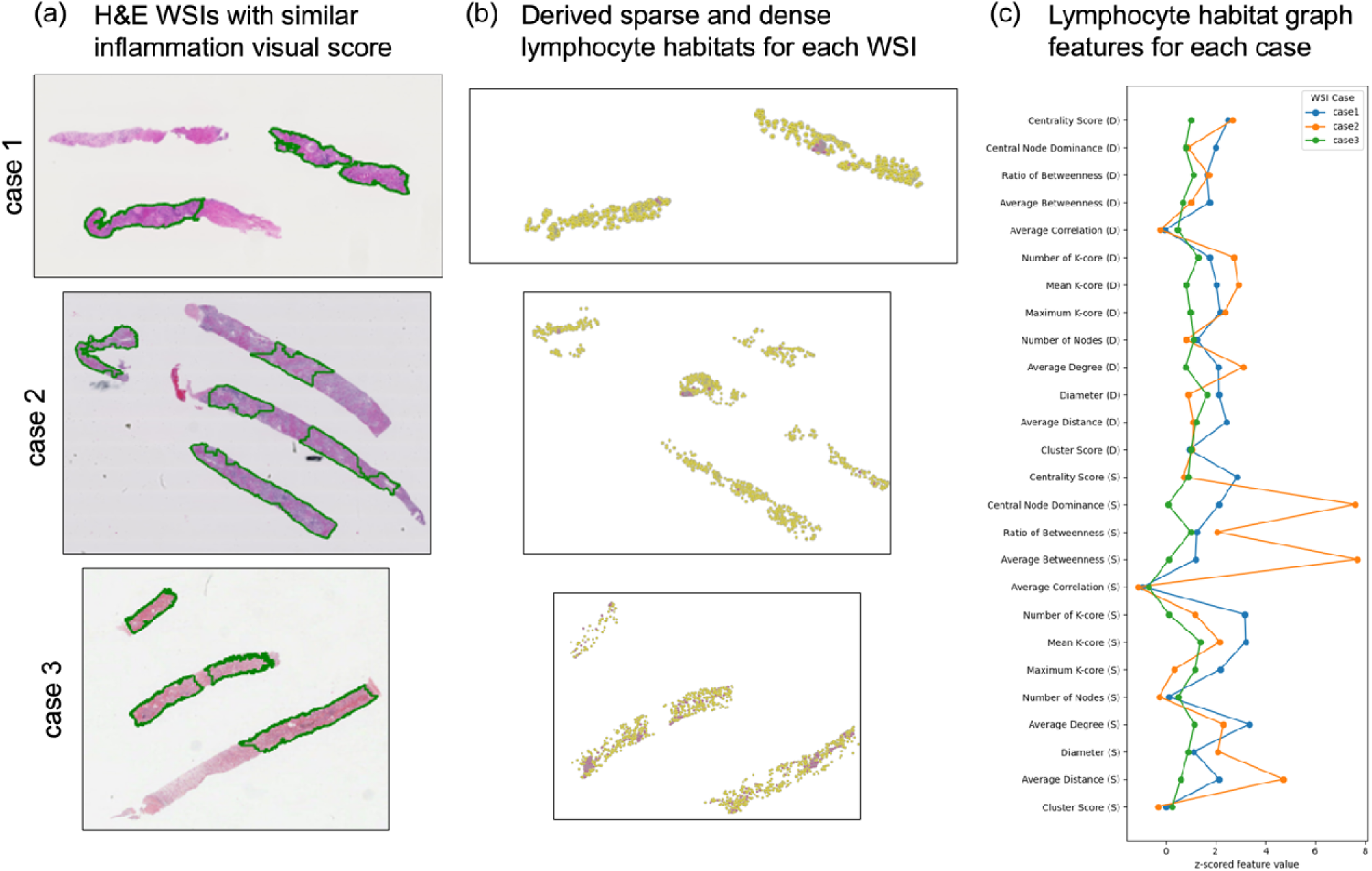
Example WSI case with similar semiquantitative metric of interstitial inflammation and distinct cell graph feature values. (a) H&E WSIs from cases with similar semiquantitative score for inflammation (green contours indicate cortex area). (b) The inflammation cell graphs with dense (pink) and sparse (yellow) habitats. (c) The cell graph feature values from dense and sparse habitats are different across cases with similar semiquantitative score for inflammation.

## Discussion

This study introduces a novel computational approach to characterize lymphocyte topology using pathomic analysis on WSIs from FSGS/MCD kidney biopsies. By leveraging whole slide cell graphs and graph theory, we have developed a method that goes beyond traditional approaches, providing a more accurate understanding of the lymphocyte spatial distribution and its potential implications for disease outcomes in kidney pathology. We identified key topological features that may serve as new biomarkers of clinical trajectory, potentially offering new insights into the underlying mechanism of disease progression. We have also demonstrated that this approach may increase our ability to predict clinical outcome.

Traditional methods for the assessment of inflammation often rely on simple measures like a gestalt/semiquantitative score approach, cell counts, or density^5,28,29^. In contrast, our approach allows for the quantification of complex topological features such as connectivity, clustering, and centrality. This provides a richer description of the inflammatory microenvironment. (Figure 6) This is particularly important in kidney pathology, where the distribution of inflammatory cells can vary significantly across different regions. Our strategy, combining local cell graphs and global cell communities, allows for a multi-scale analysis of inflammation: local graphs capture fine-grained interactions between cells, while the clustering into global communities provide a broader view of inflammatory patterns across the entire tissue section.

We also demonstrated the clinical value of this novel approach. The univariate Cox regression analysis indicates that individual lymphocyte-related signals, including some lymphocyte cell graph features, can have significant association with disease progression. However, such association is not maintained after adjustment for clinical and demographic variables, for most graph topology features, and it is maintained only for the graph feature “*average_degree* in dense habitats”. This suggests that (i) the presence of dense habitats (dense inflammation) appears to be more relevant than the presence of sparse inflammation, and (ii) spatial characteristics have potential for describing inflammation activities across the entire cortex and patterns, but more complex modeling is needed to utilize all the extracted information to test their clinical relevance.

Notably, the multivariate Cox models containing clinical/demographic parameters and graph topology features combined showed good predictive capability, but only under specific circumstances. Since we applied this computational framework across multi- and multi consortia datasets of FSGS/MCD, we conducted 2 experiments with different approaches for data partitioning. The data partition with training on one dataset (NEPTUNE) and validating on the other dataset (CureGN) showed validation performance drop. The initial low validation score on the CureGN dataset when training only on NEPTUNE dataset suggests significant differences between the two datasets, despite both enrolling adult and pediatric participants in North America with a diagnosis of FSGS or MCD. However, there was an improvement in validation scores as CureGN data was gradually added to the training se.t. The optimal mix for generalization from this NEPTUNE to CureGN dataset appeared to be when 33% of CureGN cases were included, suggesting that a balanced representation of different cohorts may be crucial for developing widely applicable predictive models.

The feature selection analysis from the multivariate Cox model (NEPTUNE+CureGN) revealed that while clinical, demographic, and visual descriptor scoring parameters (i.e., disease cohort diagnosis, interstitial fibrosis, etc.) remain strong predictors, selected graph features extracted from both the dense and sparse habitats also have predictive power. For example, in the multivariate prediction model, we introduced a non-linear feature interaction term - the product of *max_kcore* and *num_kcore* - which quantifies regional clustering strength across the tissue microenvironment. The combination of *max_kcore * num_kcore* in sparse habitats was selected frequently and was associated with a trend of decreasing risk of disease progression, indicating that (i) information extracted from sparse inflammation may have clinical relevance, (ii) the graph characteristics can be combined to better capture the lymphocyte distribution, and (iii) the number of clustered lymphocytes within the sparse habitats and a greater extent of sparse habitats compared to dense habitats is indicative of mild inflammation. Conversely, the *average_degree* in dense habitats showed a trend of increasing risk of disease progression. In other words, when there was increased connectivity among lymphocyte cells in dense habitats, this indicated a more severe inflammatory condition with patients having a higher risk of disease progression. These findings suggest that both the regional spatial clustering pattern and overall connectivity density of lymphocyte communities may provide quantitative insights into the spatial dynamics of immune cell communities that may influence disease progression.

While our study presents a novel approach to characterizing lymphocyte topology in glomerular diseases, it is important to acknowledge limitations and areas for future research. (i) although our study leveraged data from two kidney disease consortia (NEPTUNE and CureGN), the sample size remains relatively small for a complex computational model. A larger, more diverse dataset would be crucial for robust validation of our findings and to ensure generalizability across different patient populations and disease subtypes. (ii) Our current approach uses mean values of topological features from local habitats to describe the global dense and sparse habit characteristics. While this method provides valuable insights, it may simplify the complex spatial arrangements of lymphocytes. Future work could explore more sophisticated aggregation methods to identify the most informative way to summarize local features at the global level. (iii) inflammatory cells other than lymphocytes may participate in the inflammatory process and their presence, topology, and spatial interaction with other normal or abnormal kidney functional tissue units (glomeruli, tubules, arteries, peritubular capillaries, and interstitial fractional space) may have clinical relevance. Future work will focus on adapting this approach to analyze the spatial relationships between different cell types, and between inflammatory cells and kidney functional tissue units, to provide a more comprehensive view of the inflammatory landscape.

In conclusion, the use of graph theory and community detection algorithms opens to the possibility of identifying novel inflammatory patterns that may not be apparent through visual inspection or traditional quantitative and semiquantitative methods. Our study underscores the potential of integrating traditional clinical markers with novel graph topology (pathomic) features for enhanced prognostic modeling in glomerular diseases. Further work is needed to translate these complex topological features into biologically plausible signatures and clinically interpretable metrics that can ultimately guide decision-making and targeted therapies.

## Data Availability

All data produced in the present study are available upon reasonable request to the authors

## ACKNOWLEDGMENT

Research reported in this publication was supported by

1. The National Institute of Health (NIH) under the following awards: (i) National Institute of Diabetes and Digestive and Kidney Diseases (NIDDK) under the award number 2R01DK118431-04; and (ii) the National Library of Medicine (NLM) under award numbers R01LM013864.
2. The Nephrotic Syndrome Study Network (NEPTUNE) is part of the Rare Diseases Clinical Research Network (RDCRN), which is funded by the National Institutes of Health (NIH) and led by the National Center for Advancing Translational Sciences (NCATS) through its Division of Rare Diseases Research Innovation (DRDRI). NEPTUNE is funded under grant number U54DK083912 as a collaboration between NCATS and the National Institute of Diabetes and Digestive and Kidney Diseases (NIDDK). Additional funding and/or programmatic support is provided by the University of Michigan, NephCure Kidney International, Alport Syndrome Foundation, and the Halpin Foundation. RDCRN consortia are supported by the RDCRN Data Management and Coordinating Center (DMCC), funded by NCATS and the National Institute of Neurological Disorders and Stroke (NINDS) under U2CTR002818.
3. Funding for the CureGN consortium is provided by U24DK100845 (formerly UM1DK100845), U01DK100846 (formerly UM1DK100846), U01DK100876 (formerly UM1DK100876), U01DK100866 (formerly UM1DK100866), and U01DK100867 (formerly UM1DK100867) from the NIDDK/NIH. Dates of funding for first phase of CureGN was 9/16/2013-5/31/2019.
4. Patient Recruitment is supported by NephCure.
5. Additional support was also provided by NephCure and the Henry E. Haller, Jr. Foundation.

## NEPTUNE Collaborating Sites

*Atrium Health Levine Children’s Hospital, Charlotte, SC*: Susan Massengill*, Layla Lo^#^ *Cleveland Clinic, Cleveland, OH*: Katherine Dell*, John O’Toole*, John Sedor**, Victoria Grange^#^ *Children’s Hospital, Denver, CO:* Bradley Dixon*, Nathan Rogers^#^

*Children’s Hospital, Los Angeles, CA*: Rachel Lestz*, Natalie Esquivias^#^

*Children’s Mercy Hospital, Kansas City, MO*: Tarak Srivastava*, Kelsey Markus^#^ *Cohen Children’s Hospital, New Hyde Park, NY*: Christine Sethna*, Suzanne Vento^#^ *Columbia University, New York, NY:* Pietro Canetta*

*Duke University Medical Center, Durham, NC:* Opeyemi Olabisi*, Rasheed Gbadegesin**, Maurice Smith^#^

*Emory University, Atlanta, GA:* Laurence Greenbaum*, Chia-shi Wang*, Chris Fan^#^

*The Lundquist Institute, Torrance, CA:* Sharon Adler*, Janine LaPage^#^

*John H Stroger Cook County Hospital, Chicago, IL:* Amatur Amarah*

*Johns Hopkins Medicine, Baltimore, MD:* Meredith Atkinson*, Ryan Hutson^#^

*Mayo Clinic, Rochester, MN:* John Lieske, Marie Hogan, Fernando Fervenza

*Medical University of South Carolina, Charleston, SC:* David Selewski*, Cheryl Alston^#^ *Montefiore Medical Center, Bronx, NY:* Kim Reidy*, Michael Ross*, Frederick Kaskel**, Patricia Flynn^#^

*New York University Medical Center, New York, NY:* Laura Malaga-Dieguez*, Olga Zhdanova**, Laura Jane Pehrson^#^, Melanie Miranda^#^

*The Ohio State University College of Medicine, Columbus, OH*: Salem Almaani*, Laci Roberts^#^ *Riley Children’s Hospital of Indiana University, Indianapolis, IN:* Myda Khalid*, Veronica Servin^#^ *Stanford University, Stanford, CA:* Richard Lafayette*, Elizabeth Chen^#^

*Temple University, Philadelphia, PA:* Iris Lee**

*Texas Children’s Hospital at Baylor College of Medicine, Houston, TX*: Shweta Shah*, Thinh Phan^#^

*University Health Network Toronto:* Heather Reich*, Michelle Hladunewich**, Paul Ling^#^, Martin Romano^#^

*University of California at San Diego, San Diego, CA:* Ambarish Athavale*, Caitlin Carter*, Kristin Zeeb^#^

*University of California at San Francisco, San Francisco, CA*: Paul Brakeman*, Daniel Schrader *University of Colorado Anschutz Medical Campus, Aurora, CO*: James Dylewski* Nathan Rogers^#^

*University of Kansas Medical Center, Kansas City, KS*: Ellen McCarthy*, Catherine Creed^#^

*University of Miami, Miami, FL:* Alessia Fornoni*, Miguel Bandes^#^

*University of Michigan, Ann Arbor, MI:* Matthias Kretzler*, Laura Mariani*, Zubin Modi*, Amanda Williams^#^, Roxy Ni^#^

*University of Minnesota, Minneapolis, MN:* Patrick Nachman*, Michelle Rheault*, Ariel Langenberger^#^, Brady Wallner^#^

*University of North Carolina, Chapel Hill, NC:* Vimal Derebail*, Keisha Gibson*, Anne Froment^#^, Sharia Warren^#^

*University of Pennsylvania, Philadelphia, PA:* Lawrence Holzman*, Kevin Meyers**, Krishna Kallem^#^, Arielle Swenson^#^

*University of Texas San Antonio, San Antonio, TX*: Samin Sharma**

*University of Texas Southwestern, Dallas, TX:* Elizabeth Roehm*, Kamalanathan Sambandam**, Elizabeth Brown**

*University of Washington, Seattle, WA:* Ashley Jefferson*, Sangeeta Hingorani**, Katherine Tuttle^**§^, Linda Manahan ^#^, Emily Pao^#^, Kelli Kuykendall^§^

*Wake Forest University Baptist Health, Winston-Salem, NC:* Jen Jar Lin**

*Washington University in St. Louis, St. Louis, MO*: Brian Stotter*, Joseph Dumayas^#^

**Data Analysis and Coordinating Center:** *University of Michigan:* Matthias Kretzler*, Brenda Gillespie**, Laura Mariani**, Zubin Modi**, Eloise Salmon**, Howard Trachtman**, Tina Mainieri, Michael Arbit, Hailey Desmond, Sean Eddy, Damian Fermin, Wenjun Ju, Maria Larkina, Chrysta Lienczewski, Rebecca Scherr, Jonathan Troost, Amanda Williams, Yan Zhai;; *Cleveland Clinic:* Crystal Gadegbeku**, John Sedor**, *Duke University:* Laura Barisoni**; *Harvard University:* Matthew G Sampson**; *Northwestern University:* Abigail Smith**; *University of Pennsylvania:* Lawrence Holzman**, Jarcy Zee**

**Digital Pathology Committee:** Carmen Avila-Casado *(University Health Network)*, Serena Bagnasco *(Johns Hopkins University)*, Lihong Bu *(Mayo Clinic)*, Shelley Caltharp *(Emory University)*, Clarissa Cassol *(Arkana)*, Dawit Demeke *(University of Michigan)*, Brenda Gillespie *(University of Michigan)*, Jared Hassler *(Temple University)*, Leal Herlitz *(Cleveland Clinic)*, Stephen Hewitt *(National Cancer Institute)*, Jeff Hodgin *(University of Michigan)*, Danni Holanda *(Arkana)*, Neeraja Kambham *(Stanford University)*, Kevin Lemley, Laura Mariani *(University of Michigan)*, Nidia Messias *(Washington University)*, Alexei Mikhailov *(Wake Forest)*, Vanessa Moreno *(University of North Carolina)*, Behzad Najafian *(University of Washington)*, Matthew Palmer *(University of Pennsylvania)*, Avi Rosenberg *(Johns Hopkins University)*, Virginie Royal *(University of Montreal)*, Miroslav Sekulik *(Columbia University)*, Barry Stokes *(Columbia University)*, David Thomas *(Duke University)*, Ming Wu *(University of New York)*, Michifumi Yamashita *(Cedar Sinai)*, Hong Yin *(Emory University)*, Jarcy Zee *(University of Pennsylvania)*, Yiqin Zuo *(University of Miami)*. Co-Chairs: Laura Barisoni *(Duke University)*, Cynthia Nast *(Cedar Sinai)*.

**CureGN Collaborators**

The CureGN Consortium members listed below, from within the four Participating Clinical Center networks and Data Coordinating Center, are acknowledged by the authors as Collaborators.

**CureGN Principal Investigators; *CureGN Site Principal Investigators; ^+^CureGN Pathologists,

^#^CureGN Lead Coordinators.

CureGN Participating Clinical Centers (PCC) through Columbia University:

*Columbia University, New York, NY, US*: Gerald Appel, Revekka Babayev, Ibrahim Batal

^+^, Andrew Bomback**, Pietro Canetta, Brenda Chan, Vivette Denise D’Agati ^+^, Samitri Dogra, Hilda Fernandez, Gabriele Gaggero^+^, Ali Gharavi**, William Hines, Syed Ali Husain, Krzysztof Kiryluk**, Satoru Kudose ^+^, Fangming Lin, Victoria Kolupaeva#, Maddalena Marasa, Glen Markowitz ^+^, Mariela Naarro-Torres, Hila Milo Rasouly, Sumit Mohan, Nicola Mongera, Jordan Nestor, Thomas Nickolas, Jai Radhakrishnan, Maya Rao, Maya Sabatello, Simone Sanna-Cherchi, Dominick Santoriello^+^, Miroslav Sekulic ^+^, Shayan Shirazian, Michael Barry Stokes^+^, Natalie Uy, Natalie Vena, Benjamin Wooden *University of Warsaw, Warszawa, Poland:* Bartosz Foroncewicz, Natalia Krata, Barbara Moszczuk, Krzysztof Mucha*, Agnieszka Perkowska-Ptasińska, Elżbieta Ryszkowska *IRCCS Giannina Gaslini, Genoa, Italy:* Gian Marco Ghiggeri*, Francesca Lugani, Valerio Vellone^+^

CureGN Participating Clinical Centers (PCC) through the Pediatric Nephrology Research Consortium:

*Children’s Hospital of Michigan, Detroit, MI, USA*: Rossana Baracco, Amrish Jain* *Children’s Hospital of New Orleans/ LSU Health, New Orleans, LA, USA*: Diego Aviles* *Children’s Mercy Hospital, Kansas City, MO, USA*: Tarak Srivastava*, Alexander Katz^+^ *Children’s National Medical Center, Wmcgashington DC, USA*: Sun-Young Ahn* *Cincinnati Children’s Hospital Cincinnati, OH, USA*: Prasad Devarajan, Elif Erkan*, Donna Claes, Hillarey Stone

*Connecticut Children’s Medical Center, Hartford, CT, USA*: Sherene Mason* *East Carolina University Brody School of Medicine, Greenville, NC, USA*: Liliana Gomez-Mendez*

*Emory University, Atlanta, GA, USA*: Larry Greenbaum**, Chia-shi Wang, Hong (Julie) Yin^+^

*Helen DeVos Children’s Hospital, Grand Rapids, MI, USA*: Goebel Jens*, Julia Steinke

*Levine Children’s Hospital/Atrium Health, Charlotte, NC, USA*: Donald Weaver*

*Lurie Children’s Hospital, Chicago IL, USA*: Jerome Lane*

*Mayo Clinic, Rochester, MN, USA*: Carl Cramer*

*Medical College of Wisconsin, Milwaukee, WI, USA*: Cindy Pan, Neil Paloian, Rajasree Sreedharan*

*Medical University of South Carolina, Charleston SC, USA*: David Selewski, Katherine Twombley*, Sally Self^+^

*Nationwide Children’s Hospital, Columbus, OH, USA*: Samantha Martinek-Bundt#, Dawson Carmean#, Mary Dreher^#^, Aria Dockham^#^, Mahmoud Kallash*, John Mahan, Samantha Sharpe^#^, William Smoyer**, Laura Biederman^+^

*Oregon Health and Science University, Portland, OR, USA*: Amira Al-Uzri*, Sandra Iragorri

*Riley Children’s Hospital, Indianapolis, IN, USA*: Myda Khalid**

*Cardinal Glennon Children’s Medical Center/ St. Louis University, St. Louis, MO, USA*: Craig Belsha*

*Texas Children’s Hospital, Houston, TX, USA*: Elizabeth Onugha*, Michael Braun, AC Gomez

*Texas Tech Health Sciences Center, Amarillo, TX, USA*: Tetyana Vasylyeva*

*Children’s of Alabama, University of Alabama, Birmingham, AL, USA*: Daniel Feig*

*University of Colorado Children’s Hospital, Colorado, Aurora, CO, USA*: Melisha Hannah*

*University of Iowa Children’s Hospital, Iowa City, IA, USA*: Carla Nester*

*University of Kentucky, Lexington, KY, USA*: Aftab Chishti*

*University of Louisville, Louisville, KY, USA*: Jon Klein**

*Holtz Medical Center, University of Miami, Miami, FL, USA*: Chryso Katsoufis, Wacharee Seeherunvong*

*University of Minnesota Children’s Hospital, Minneapolis, MN, USA*: Michelle Rheault** *University of New Mexico Health Sciences Center, Albuquerque, NM, USA*: Craig Wong*

*University of Oklahoma Health Sciences Center, Oklahoma City, OK, USA*: Qassim Abid*

*University of Virginia, Charlottesville, VA, USA*: John Barcia*, Agnes Swiatecka-Urban

*University of Wisconsin, Madison, WI, USA*: Sharon Bartosh*

*Vanderbilt Children’s Hospital, Nashville TN, USA*: Tracy Hunley*

*Washington University in St. Louis, St. Louis, MO, USA*: Vikas Dharnidharka*, Brian Stotter, Joseph, Gaut ^+^

CureGN Participating Clinical Centers (PCC) through the University of North Carolina:

*Hôpital Maisonneuve-Rosemont, Montreal, Canada*: Louis-Philippe Laurin*, Virginie Royal^+^, Mathieu Latour^+^, Natlie (Natacha) Patey ^+^

*Medical University of South Carolina, Charleston, SC, USA*: Anand Achanti, Milos Budisavljevic*

*Northwestern University, Chicago, IL, USA*: Cybele Ghossein, Yonatan Peleg * *Ohio State University, Columbus, OH, USA*: Salem Almaani*, Isabelle Ayoub, Samir Parikh, Brad Rovin, Anjali Satoskar^+^

*University of Chicago, Chicago, IL, USA*: Anthony Chang^+^

*University of Alabama at Birmingham, Birmingham, AL, USA*: Huma Fatima^+^, Jan Novak, Matthew Renfrow, Dana Rizk*

*<u>University of North Carolina Kidney Center, Chapel Hill, NC, USA</u>*: Dhruti Chen, Vimal Derebail**, Ronald Falk**, Keisha Gibson, Dorey Glenn, Susan Hogan, Koyal Jain, J. Charles Jennette^+^, Amy Mottl, Caroline Poulton^#^, Monica Reynolds, Manish Kanti Saha, Nicole E. Wyatt

*Vanderbilt University, Nashville, TN, USA*: Agnes Fogo^+^, Neil Sanghani* *Virginia Commonwealth University, Richmond, VA, USA*: Jason Kidd*, Selvaraj Muthusamy^+^

CureGN Participating Clinical Centers (PCC) through the University of Pennsylvania:

*Children’s Hospital of Philadelphia, Philadelphia, PA, USA*: Michelle Denburg, Amy Kogon, Kevin Meyers*, Madhura Pradhan

*Cleveland Clinic, Cleveland, OH, CA*: Raed Bou Matar*, John O’Toole, John Sedor Cohen Children’s Medical Center, New Hyde Park, NY, USA: Christine Sethna*^, Suzanne Vento^#^

*Johns Hopkins University, Baltimore, MD, USA*: Mohamed Atta, Serena Bagnasco^+^, Alicia Neu, John Sperati*

*Lundquist Institute at Harbor-UCLA Medical Center, Torrance, CA, USA*: Sharon Adler*, Tiane Dai, Ram Dukkipati

*Mayo Clinic, Rochester, MN, USA*: Fernando Fervenza*, Sanjeev Sethi ^+^

*Montefiore Medical Center, The Bronx, New York, NY, USA*: Frederick Kaskel, Kaye Brathwaite, Kimberly Reidy*

*New York University, New York, NY, USA*: Joseph Weisstuch, Ming Wu ^+^, Olga Zhdanova

*Spokane Providence Medical Center, Spokane, WA, USA*: Katherine Tuttle* *Stanford University, Palo Alto, CA, USA*: Jill Krissberg, Richard Lafayette*, Kamal Fahmeedah, Elizabeth Talley

*Sunnybrook Health Sciences Centre, Toronto, Canada*: Michelle Hladunewich*

*The Hospital for Sick Children, Toronto, Canada*: Rulan Parekh*

*University Health Network, Toronto, Canada*: Carmen Avila-Casado^+^, Daniel Cattran*, Reich Heather, Philip Boll

*University of Miami, Miami, FL, USA*: Yelena Drexler, Alessia Fornoni*

*University of Michigan, Ann Arbor, MI, USA*: Jeffrey Hodgin^+^, Andrea Oliverio* *University of Pennsylvania, Philadelphia, PA, USA*: Jon Hogan, Lawrence Holzman**, Matthew Palmer ^+^, Gaia Coppock

*University of Pittsburgh School of Medicine, Pittsburgh, PA, USA*: Michael Mortiz* *University of Washington, Seattle, WA, USA*: Charles Alpers^+^, J. Ashley Jefferson* *UT Southwestern, Dallas, TX, USA*: Kamal Sambandam*, Bethany Roehm

## Data Coordinating Center (DCC)

*Cedar Sinai Medical Center, Los Angeles, CA, USA*: Cynthia Nast^+^, Jean Hou^+^

*Duke University, Durham, NC, USA*: Laura Barisoni *Cleveland Clinic, Cleveland, OH, USA:* Crystal Gadegbeku** *Northwestern University, Chicago, IL, USA:* Abigail Smith**

*University of Michigan, Ann Arbor, MI, USA*: Brenda Gillespie, Bruce Robinson, Matthias Kretzler, Zubin Modi, Laura Mariani**

**Steering Committee Chair:** Lisa M. Guay-Woodford, Children’s Hospital of Pennsylvania, Philadelphia, PA, USA

