## Supplemental Table 1 for "Computational characterization of lymphocyte topology on whole slide images of glomerular diseases"

**Supplemental Table 1. Graph feature list.**

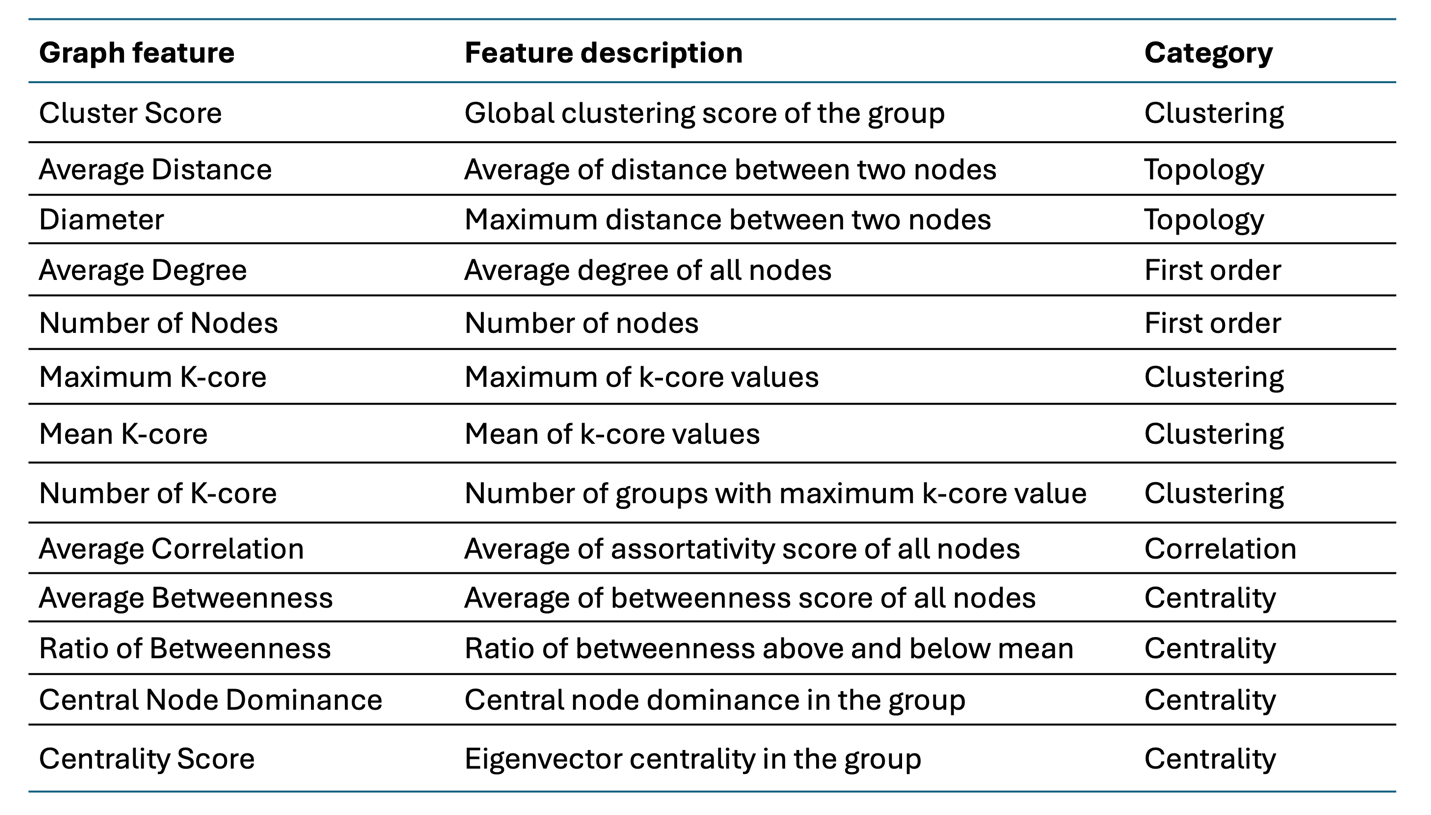

**Cluster Score**: Global clustering score measures how tightly interconnected nodes are within groups. It quantifies the tendency of nodes to form densely connected clusters by calculating the ratio of actual triangles (three fully connected nodes) to potential triangles in the network.

**Average Distance:** The mean of shortest path lengths between all pairs of nodes in the network. A shorter average distance indicates nodes are generally closer and information can flow more efficiently through the network.

**Diameter:** The maximum shortest path length between any two nodes in the network. This represents the "longest" distance you'd need to traverse to get from one node to another, indicating the network's overall span.

**Average Degree:** The mean number of connections (edges) that nodes have in the network. Higher average degree suggests nodes are more interconnected, while lower values indicate sparser connectivity.

**Number of Nodes:** Total count of vertices/points in the network. This fundamental measure determines the network's size and impacts computational complexity of various analyses.

**Maximum K-core:** The highest k value for which a k-core exists in the network. A k-core is a maximal subgraph where all vertices have degree at least k, indicating the most densely connected part of the network.

**Mean K-core:** Average k-core value across all nodes. This indicates the typical level of core membership for nodes, helping understand the overall cohesiveness of the network structure.

**Number of K-core:** Count of distinct groups that achieve the maximum k-core value. This shows how many highly connected subcommunities exist at the network's densest level.

**Average Correlation:** Mean assortativity score measuring the tendency of nodes to connect to other nodes with similar characteristics (like degree). Higher values indicate stronger homophily in connections.

**Average Betweenness:** Mean betweenness centrality across all nodes, indicating how often nodes act as bridges along shortest paths between other nodes. Higher values suggest more nodes play important intermediary roles.

**Ratio of Betweenness:** Comparison of betweenness centrality values above and below the mean, helping identify if bridging roles are evenly distributed or concentrated in certain nodes.

**Central Node Dominance:** Measures how much influence the most central nodes have over the network structure, indicating whether control/influence is concentrated or distributed.

**Centrality Score:** Eigenvector centrality measurement for the group, which reveals influence patterns in the network. Eigenvector centrality is a measure of influence that considers both direct and indirect connections in a network, working on the principle that connections to high-scoring nodes contribute more to a node's score than equal connections to low-scoring nodes.
